# Cost-effectiveness of digital therapeutics for essential hypertension

**DOI:** 10.1101/2022.03.02.22271583

**Authors:** Akihiro Nomura, Tomoyuki Tanigawa, Kazuomi Kario, Ataru Igarashi

## Abstract

**Background:** Hypertension increases the risk of cardiovascular and other diseases. Lifestyle modification is a significant component of nonpharmacological treatments for hypertension. We previously reported the clinical efficacy of digital therapeutics (DTx) in the HERB-DH1 trial. However, there is still a lack of cost-effectiveness assessments evaluating the impact of prescription DTx. This study aimed to analyze the cost-effectiveness of using prescription DTx in treating hypertension.

**Methods:** We developed a monthly cycle Markov model and conducted Monte Carlo simulations using the HERB-DH1 trial data to investigate quality-adjusted life-years (QALYs) and the cost of DTx for hypertension plus guideline-based lifestyle modification consultation treatment as usual (TAU), comparing DTx+TAU and TAU-only groups with a lifetime horizon. The model inputs were obtained from the HERB-DH1 trial, published or publicly available data, and expert assumptions. The incremental cost-effectiveness ratio (ICER) per QALY was used as the benchmark for cost-effectiveness. We performed probabilistic sensitivity analyses (PSAs) using the Monte Carlo simulation with 2 million sets.

**Results:** The DTx+TAU strategy produced 18.778 QALY and was associated with ¥3,924,075 ($34,122) expected costs, compared with 18.686 QALY and ¥3,813,358 ($33,160) generated by the TAU-only strategy over a lifetime horizon, resulting in an ICER of ¥1,199,880 ($10,434)/QALY gained for DTx+TAU. The monthly cost and attrition rate of DTx for hypertension have a significant impact on ICERs. In the PSA, the probability of the DTx arm being a cost-effective option was 87.8% at a threshold value of ¥5 million ($43,478)/QALY gained.

**Conclusions:** The DTx+TAU strategy was more cost-effective than the TAU-only strategy.

## Introduction

Hypertension or elevated blood pressure (BP) is a serious health condition that significantly increases cardiovascular diseases (CVD), heart failure (HF), atrial fibrillation (AFib), renal dysfunction, and other disorders.^1^ Hypertension affects 1.28 billion people and is a leading modifiable risk factor attributing mortality worldwide.^1, 2^ In Japan, there were more than 43 million patients with hypertension, having one of the highest prevalence among the Organization for Economic Co-operation and Development countries.^3^ Hypertension is attributed to cerebral and cardiovascular deaths, accounting for 50% of all CVD deaths, 59% of coronary artery disease (CAD) deaths, and 52% of stroke deaths.^4^ Also, the total medical cost of these CVDs has reached nearly $100 billion in Japan.^5^ Therefore, managing hypertension is one of the most critical issues for preventing CVDs and reducing medical costs in Japan. For this purpose, Health Japan 21 (the second edition) aims to reduce the average systolic BP (SBP) of the Japanese population by 4 mmHg within 10 years by raising awareness and promoting lifestyle modification by a healthy diet, more physical activity, and reducing alcohol consumption at the national level.^6^

Lifestyle modification is a major component of non-pharmacological treatment for hypertension, which is as essential as antihypertensive pharmacological treatment.^7^ In principle, hypertensive patients at low risk for CVD should modify their lifestyle first, and antihypertensive drug therapy is initiated at an appropriate time according to the patients’ future CVD risks. Lifestyle modification plays a vital role in lowering blood pressure because of its antihypertensive and synergistic effects along with antihypertensive drugs.^8, 9^ However, the physician’s limited outpatient time is insufficient to provide guideline-based hypertension management, including lifestyle modification suited for each patient’s diverse lifestyle.^10^ As a result, lifestyle modification has not achieved its full effect in reducing BP.

To maximize the potential treatment effect of lifestyle modification for hypertension, we developed a prescription digital therapeutics (DTx) for hypertension, the HERB system.^11^ The HERB system consists of a HERB Mobile smartphone app for patients and a web-based patient management software for primary doctors.^11^ We previously reported the HERB-DH1 phase III randomized controlled trial (RCT) results of the DTx for essential hypertension. In this trial, we randomly assigned 390 essential hypertension patients aged ≤65 years who were not taking antihypertensive medication to the DTx intervention group or the control group. The results showed that the primary outcome of the 24-h SBP was significantly lower in the DTx intervention group than that in the control group.^12^

Although the HERB-DH1 trial demonstrated the clinical efficacy of DTx, there is still a lack of cost-effectiveness assessments evaluating the impact of the prescription DTx for hypertension on healthcare costs along with clinical efficacy. The cost-effectiveness of DTx for opioid use disorder or that for low back pain were recently evaluated using the RCT results.^13–15^ However, there is no study assessing the cost-effectiveness of prescription DTx for hypertension using the RCT data. Exploring the appropriate DTx pricing determined by cost-effectiveness analysis is crucial in countries including Japan, whose DTx costs could be reimbursed by public health insurance.

Here, we aimed to analyze the cost-effectiveness of using prescription DTx in treating hypertension using the HERB-DH1 phase III RCT results.

## Methods

We developed an economic model to assess the cost and effectiveness of DTx for essential hypertension. The model investigated quality-adjusted life-years (QALYs) and the cost of DTx for hypertension plus guideline-based lifestyle modification consultation treatment as usual (TAU) (DTx+TAU) vs. TAU only with a lifetime horizon and a 1-month cycle length. We constructed the model from the perspective of a public health care payer. We compared the cost-effectiveness of DTx+TAU with TAU alone for the treatment of CAD, stroke, HF, and AFib. We used the parameters of the HERB-DH1 phase III clinical trial^12^ as inputs. The HERB-DH1 trial is an open-label RCT of HERB prescription DTx in 390 patients with essential hypertension. At 12 weeks, DTx+TAU intervention group exhibited a significant reduction of SBP compared with the TAU-only control group (−10.6 mmHg in DTx+TAU group vs −6.2 mmHg in the TAU-only control group by a home BP measurement). The present study evaluated outcomes as incremental cost-effectiveness ratio (ICER) per QALY gained. The model inputs were obtained from the HERB-DH1 trial data, published academic papers, publicly available data, and expert assumptions. We conducted the analyses according to the Consolidated Health Economic Evaluation Reporting Standards (CHEERS) statement and the Ethics Guidelines for Medical and Biological Research Involving Human Subjects in Japan. The authors have the right to publish, regardless of the outcome. The Institutional Review Board approved the study protocol at Kanazawa University, Japan.

### Model structure and clinical pathway

We developed a monthly cycle Markov model and conducted Monte Carlo simulations to estimate the efficacy of DTx+TAU (**Figure 1**). A Monte Carlo simulation, with 1,000,000 iterations, was performed to reflect the variation in patient characteristics, such as age and treatment effects, mainly derived from those of enrollees for the clinical trial. The model consisted of patients who received DTx+TAU and those who received TAU only. We modelled three different health statuses as follows: “stay” (healthy without any acute complications), “acute complications*”* (suffering any acute complications regarding hypertension), and “death.” Acute complications included CAD, stroke, HF, and AFib. We also considered natural deaths in the model. Hypertensive patients who avoid natural death can stay in the same acute complication-free status “stay” or experience acute complications of CAD, stroke, HF, or AFib. In this model, a patient with one of the acute complications must maintain the “acute complication” status or die directly from the complication for 12 months, then automatically recover and switch from the “acute complication” status to the “stay” status with a post-complication (chronic) tag [return to the “stay” status but had slightly higher mortality rates than natural deaths in patients after having either of CAD, stroke, and HF]. We did not consider a patient with more than one complication within 12 months of the latest occurrence of acute complications.

**Figure 1.**
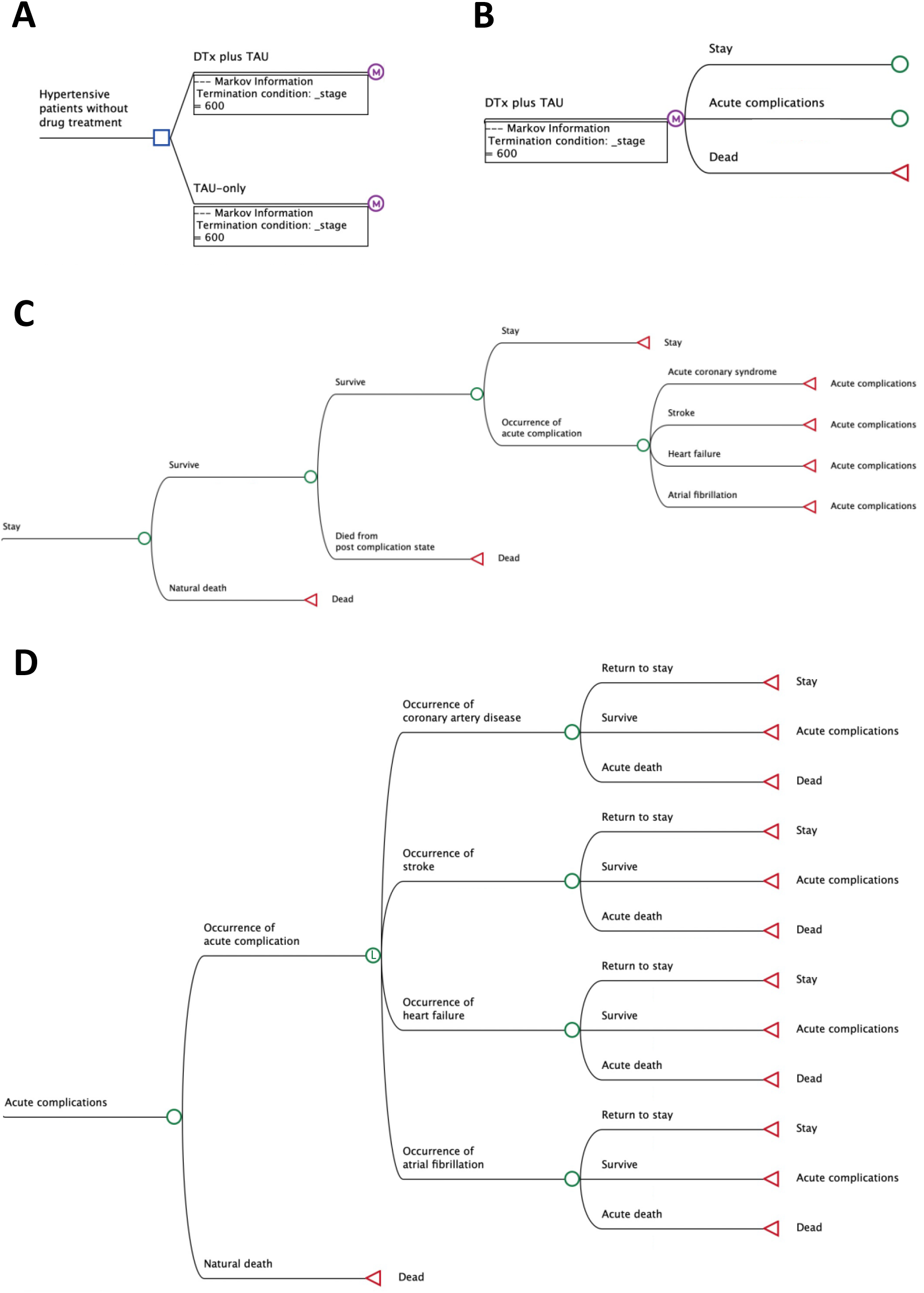
Structure of the simulation model. A) Decision tree. B) Structure of Markov model. C) Structure of Stay (no acute complications) module. D) Structure of acute complications module.

### Time horizon

We set lifetime years (50 years) as the time horizon for our study. We also conducted scenario analyses to slide the time horizon to 20, 30, and 40 years.

### Model population and intervention

The study population was derived from the HERB-DH1 trial. They were grade I or II hypertensive patients without antihypertensive drug treatment, with a mean age of 52 years, 20% were female, 16% were current smokers, and the proportion of dyslipidemia and diabetes was 50% and 7%, respectively. The average SBPs was 145 mmHg in the 24-h SBP by ambulatory BP monitoring and 148 mmHg in the morning home SBP by home BP monitoring. This model used log-normalized baseline home SBP distribution from HERB-DH1 trial data (**Table 1**). Additionally, we modelled annual SBP increase in both groups by 0.5 mmHg from the national survey.^7^

**Table 1.** Model inputs.

| Data | Input | Low* | High* | Source |
| --- | --- | --- | --- | --- |
| Age | 52.3 years | - | - | Kario, 2021. |
| Baseline SBP | 148 mmHg | - | - | Kario, 2021. |
| Annual BP increase | 0.5 mmHg | 0.39 | 0.65 | Umemura, 2019 |
| Annual mortality rates by complications |  |  |  |  |
| Coronary artery disease |  |  |  |  |
| First 1 month after event onset | 5.0% / month | 0 | 5.0 | Komiyama, 2018. |
| > 1 month (post-CAD) | 0.98% | 0.93 | 1.74 | Goto, 2011. |
| Stroke |  |  |  |  |
| First 1 month after event onset | 12.9% / month | 9.7 | 16.1 | Takashima, 2017. |
| > 1 month (post-Stroke) | 1.29% | - | - | Goto, 2011. |
| Heart failure |  |  |  |  |
| First 12 months after event onset | 11.5% | 10.1 | 13.0 | Shiraishi, 2018. |
| >13 months (post-HF) | 2.5% | - | - | Tsuji, 2017. |
| Atrial fibrillation |  |  |  |  |
| First 12 months after event onset | 1.83% | 1.28 | 2.56 | Goto, 2011. |
| >13 months (post-AFib) | Same as the natural mortality rate |  |  | - |
| Annual incidence rates of each complication among hypertension patients |  |  |  |  |
| Coronary artery disease | 0.460% | 0.345 | 0.575 | Kaneko, 2021. Turin, 2010. |
| Stroke | 1.592% | 1.194 | 1.990 | Kaneko, 2021. Turin, 2016. |
| Heart failure | 0.889% | - | - | Kaneko, 2021. Fujimoto 2021. |
| Atrial fibrillation | 0.232% | - | - | Kaneko, 2021. |
| Annual costs |  |  |  |  |
| DTx for hypertension | ¥120,000 (\$1,043) | 60,000 | 240,000 | Assumption |
| Acute phase (First 12 months after event onset) |  |  |  |  |
| Coronary artery disease | ¥2,156,290 (\$18,750) | 1,617,218 | 2,695,362 | Kamae, 2015. |
| Stroke | ¥1,440,107 (\$12,523) | 1,080,080 | 1,800,134 | Kamae, 2015. |
| Heart failure | ¥770,428 (\$6,699) | 577,821 | 963,035 | Mizuno, 2017. |
| Atrial fibrillation | ¥120,000 (\$1,043) | 90,000 | 150,000 | Kamae, 2018. |
| Post-complication phase (>12 months) |  |  |  |  |
| Coronary artery disease | ¥495,600 (\$4,310) | 459,996 | 570,000 | Kodera, 2019 |
| Stroke | ¥318,387 (\$2,769) | 262,992 | 372,996 | Kodera, 2019 |
| Heart failure | ¥770,428 (\$6,699) | 577,821 | 963,035 | Mizuno, 2017. |
| Atrial fibrillation | ¥120,000 (\$1,043) | 90,000 | 150,000 | Kamae, 2018. |
| Health utilities |  |  |  |  |
| Normal | 0.950 | 0.760 | 1.000 | - |
| Coronary artery disease (increment) | -0.180 | -0.135 | -0.225 | Kodera, 2019. |
| Stroke (increment) | -0.157 | -0.117 | -0.196 | Hattori, 2012. |
| Heart failure (increment) | -0.101 | -0.076 | -0.126 | Clarke 2002. |
| Atrial fibrillation (increment) | -0.000 | - | - | Assumption |
| Death (increment) | -0.950 | - | - | - |
| Discount rates | 2% | 0 | 4 | Shiroiwa, 2017. |

\* Range for sensitivity analysis.
Abbreviations: AF, atrial fibrillation; BP, blood pressure; CAD, coronary artery disease; CI, confidence interval; HF, heart failure; SBP, systolic blood pressure.

The DTx+TAU group received the prescription DTx, HERB Mobile system, and lifestyle modifications to manage hypertension recommended by the Japanese Society of Hypertension (JSH) guidelines.^7^ The HERB Mobile system include the HERB Mobile smartphone app and a web-based management application for healthcare providers. The smartphone app retrieves each patient’s baseline input data, and daily home BP measurements transfer the data to the cloud server and assess them to plan a personalized lifestyle modification program to lower their BP. In addition, primary physicians can simultaneously browse their patients’ data such as BP measurements, daily activities, or progress on the proposed program using a web-based application.^11, 12^ The TAU-only group also promoted lifestyle modifications for hypertension based on the JSH guidelines by their primary physicians or themselves. We assume that the hypertensive patients in the DTx+TAU group have an average home SBP reduction of 10.6 mmHg, while those in the TAU-only group had a 6.2 mmHg reduction from the previous study.^12^ In this simulation model, we used DTx+TAU and TAU-only effect distributions derived from the HERB-DH1 trial data (**Supplemental Figure**). Since DTx adherence could decrease with time, we set the model such that 25% of the remaining users in the DTx+TAU group attrited using the DTx every 6 months.

### Mortality and complications

We obtained the annual natural death rates from the Japan Vital Statistics for 2022.^16^ the annual mortality and occurrence rates of each complication are summarized in **Table 1**. We set both acute and post-state (chronic) mortality rates for each complication.^17–21^

The occurrence rates of each complication were adjusted according to the BP categories (**Supplemental Table**).^17^ Additionally, the annual incidence rates of CAD and stroke were calibrated referring to the previously reported lifetime risks.^22, 23^ In addition, the annual incidence rate of HF was calibrated by a patient’s age (hazard ratios [HRs] of 2.5, 5.0, and 10 for the 60s, 70s, and ≥80s).^24^ Categories of normal BP, elevated BP, grade I HT, and grade II HT were defined as home SBPs of <125, 125-134, 135-144, and ≥145 mmHg, respectively.^7^ Since the cycle length of the model was 1 month, we converted the annual rates to the monthly rates to input the model.

### Costs

A list of the cost data is presented in **Table 1**. We performed an economic estimation from Japan’s public healthcare payer perspective, under which only medical costs were considered. The costs consist of treatment and hospitalization expenses, but not transportation fees or family care service costs. We set the monthly cost of HERB DTx at ¥10,000 ($86.96 US: $1 US = ¥115 as of January 2022), including the app purchases and subscription fees. The cost of each complication (CAD, stroke, HF, and AFib) was also derived from previous studies.^25–28^ We set annual cost reduction by 2% according to the Japanese guideline for cost-effectiveness analyses.^29^

### Health-related quality of life and utilities

We derived the utility values of CAD, stroke, HF, and AFib from published studies based on the European Quality of Life-5 Dimensions Questionnaire (**Table 1**).^28, 30, 31^ We calculated the QALY by multiplying the time duration in a specific health state by the utility value associated with the state. Both cost and outcome were discounted at a 2% discount rate.^29^

### Main analysis

We calculated the ICER between DTx+TAU and TAU-only strategies over the lifetime horizon using the estimated model parameters and cost and effectiveness assumptions.

### Sensitivity and scenario analyses

We tested the impact of the uncertainties around the model parameters on the overall cost-effectiveness results using one-way deterministic and probabilistic sensitivity analyses. First, we performed one-way deterministic sensitivity analyses by changing the treatment effect, cost, and utility parameters over each range between low and high values (**Table 1 and Supplemental Table**). Then, we conducted probabilistic sensitivity analyses to evaluate the sensitivity of the results to simultaneous variable changes using a set of 2 million (2,000 iterations × 1,000 individuals) simulated results with specific probability distributions of the input parameters. We assumed a Weibull distribution for age, log-normal distribution for initial SBP input, normal distributions for HRs of each complication by BP grades, mortality rates, and health utilities, beta distributions for clinical event incidence rates, and gamma distributions for costs. In addition, we drew the probability of the cost-effectiveness acceptability curve to evaluate the cost-effectiveness probability at a willingness-to-pay threshold of ¥5 million ($43,478). Additionally, we conducted two scenario analyses as follows: #1) simulating different time horizons of 20, 30, and 40 years; and #2) setting an alternative DTx app attrition rate of 10% every 6 months. All model analyses were conducted using TreeAge Pro HealthCare version 2022 R1.0 (TreeAge Software, LLC, MA, USA).

## Results

### Main analysis

The model predicted that the DTx+TAU strategy produced 18.778 QALY and was associated with ¥3,924,075 ($34,122) expected costs compared to 18.686 QALY and ¥3,813,358 ($33,160) generated by TAU-only treatment over a lifetime horizon. The introduction of DTx would increase costs by ¥110,717 ($962) and prolong QALY by 0.092. The ICER of DTx+TAU against TAU would be ¥1,199,880 ($10,434)/QALY gained, which were well below the hypothetical threshold value in Japan, or ¥5 million/QALY gained (**Table 2**).

**Table 2.** Cost-effectiveness of DTx for hypertension.

| Outcome | DTx+TAU | TAU-only | Changes |
| --- | --- | --- | --- |
| Lifetime horizon |  |  |  |
| Costs | ¥3,924,075 (\$34,122) | ¥3,813,358 (\$33,160) | ¥110,717 (\$962) |
| QALYs | 18.778 | 18.686 | 0.092 |
| ICER | ¥1,199,880 (\$10,434)/QALY | | |
| 40-year time horizon |  |  |  |
| Costs | ¥3,705,428 | ¥3,591,385 | ¥114,043 |
| QALYs | 18.483 | 18.395 | 0.089 |
| ICER | ¥1,284,159 (\$11,167)/QALY | | |
| 30-year time horizon |  |  |  |
| Costs | ¥2,996,162 | ¥2,869,894 | ¥126,268 |
| QALYs | 17.195 | 17.124 | 0.071 |
| ICER | ¥1,788,344 (\$15,551)/QALY | | |
| 20-year time horizon |  |  |  |
| Costs | ¥1,820,066 | ¥1,664,483 | ¥155,583 |
| QALYs | 13.939 | 13.903 | 0.037 |
| ICER | ¥4,235,677 (\$36,832)/QALY | | |

Abbreviations: DTx, digital therapeutics; ICER, incremental cost-effectiveness ratio; QALY, quality-adjusted life year; TAU, treatment as usual

### Sensitivity analyses

We conducted one-way deterministic and probabilistic sensitivity analyses. The results of the one-way deterministic sensitivity analyses with the tornado diagram are shown in **Figure 2**. The monthly cost of DTx, which ranged from ¥5,000 ($43.48)/month to ¥20,000 ($173.91)/month, had the largest impact on the ICER value, followed by the DTx attrition and discount rates. According to the one-way threshold analysis for the monthly DTx cost, the DTx+TAU arm became the dominant strategy when the cost was below ¥5,163 ($44.90) per month. Even the highest ICER value observed in this diagram was still below the threshold value, or ¥5 million/QALY gained.

**Figure 2.**
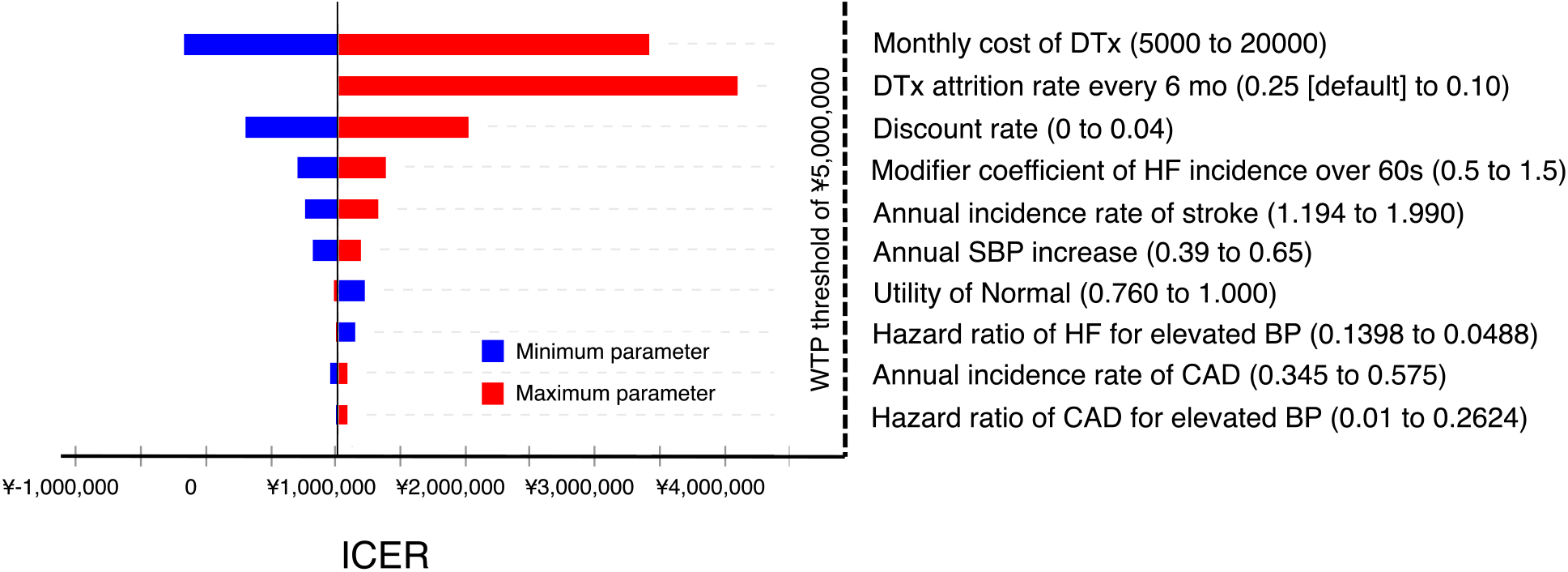
One-way sensitivity analysis results.

The results of the probabilistic sensitivity analysis are shown with incremental cost-effectiveness scatter plots (**Figure 3**) and a cost-effectiveness acceptability curve (**Figure 4**). According to these figures, the probability of dominance (DTx less costly and more effective) and cost-effectiveness (DTx more costly and more effective, the ICER was less than ¥5 million /QALY gained) were 6.5% and 81.3%, respectively.

**Figure 3.**
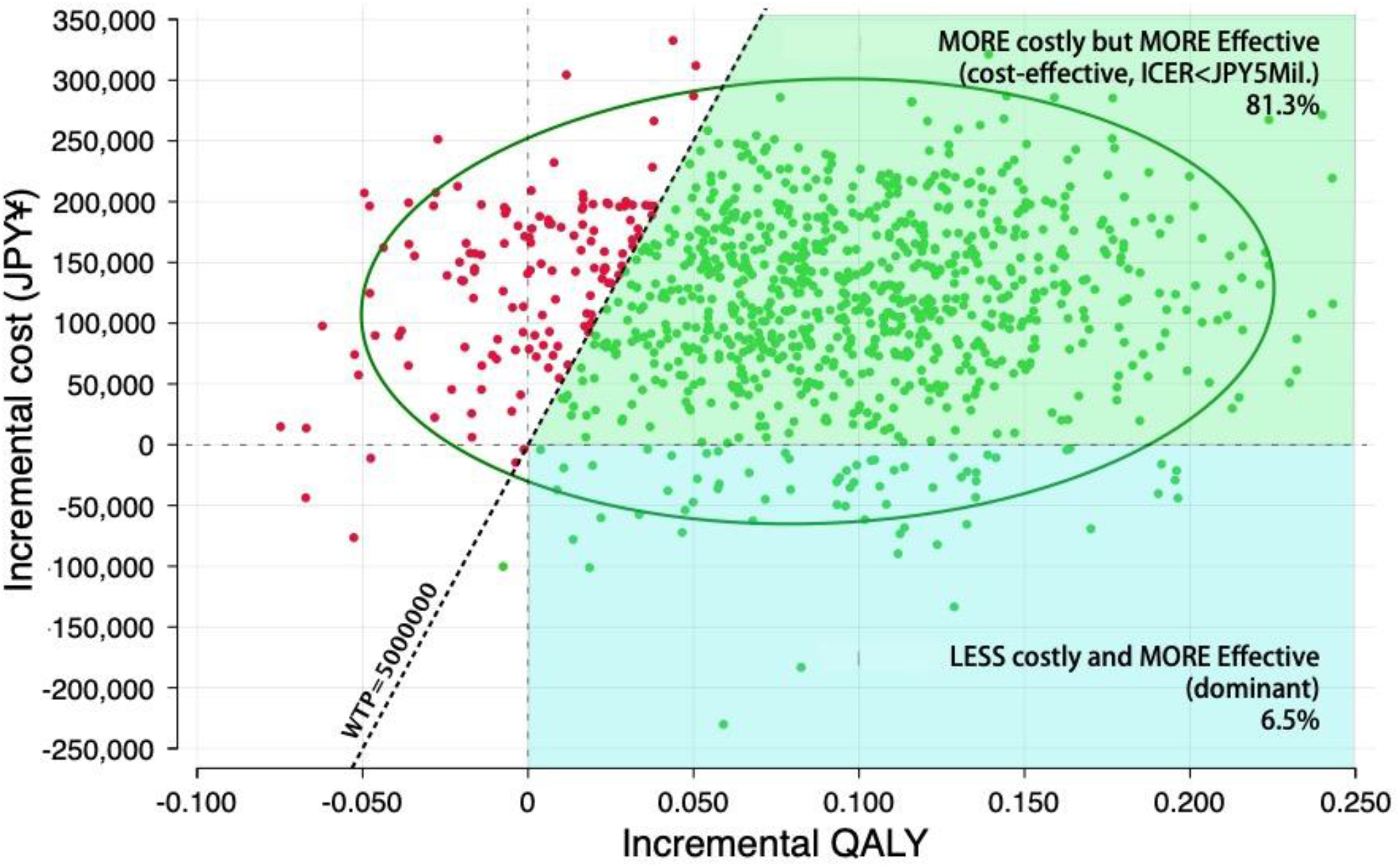
Cost-effectiveness plot.

**Figure 4.**
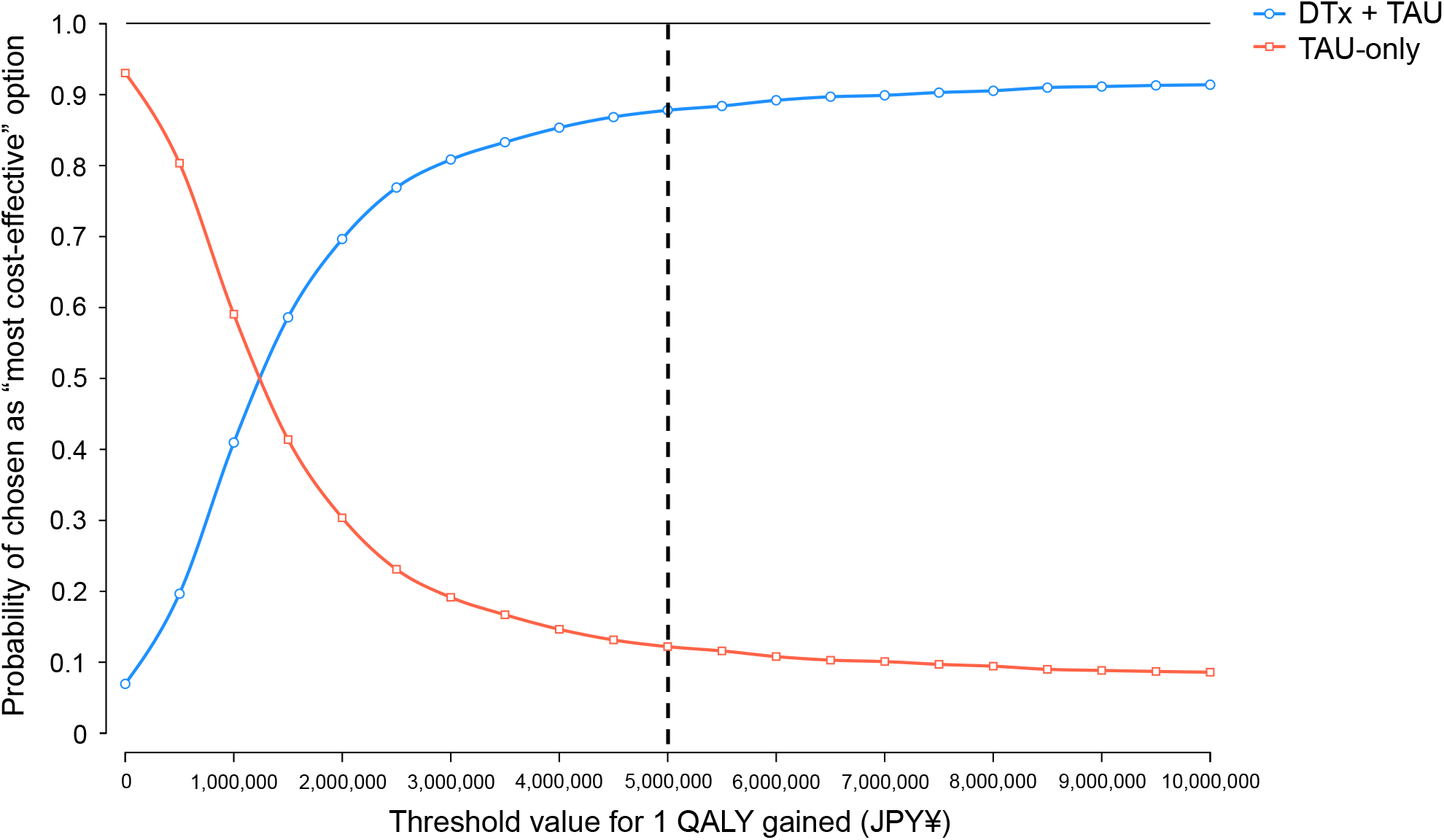
Cost-effectiveness acceptability curve in DTx+TAU and TAU-only group.

### Scenario analyses

In scenario #1, by changing the simulation duration, the ICERs for DTx+TAU treatment were inversely proportional to the extent of the time horizon (Table 2). In scenario #2, using an alternative DTx attrition rate of 10% every 6 months, ICER was reached in ¥4,330,295 ($37,655)/QALY.

## Discussion

In this study, we assessed the cost-effectiveness of prescription DTx for essential hypertension, in addition to TAU, compared to the TAU-only strategy. We found that the ICER of the DTx +TAU strategy was ¥1,199,880 ($10,434) /QALY against the TAU-only strategy in the lifetime horizon, which was well below the threshold value, or ¥5 million /QALY. Moreover, the probability of the cost being below the threshold value was 87.8% in the probabilistic sensitivity analysis. This model analysis demonstrated that DTx+TAU is cost-effective for patients with hypertension.

There are several conclusions that can be drawn from this study. First, this is the first study to evaluate the cost-effectiveness of DTx for patients with hypertension using the results of an RCT. Nordyke et al. reported the good cost-effectiveness of DTx for hypertension with person-to-person health coaching from retrospective study data over a 3 year time horizon.^32, 33^ Other than hypertension, DTx for opioid use disorder and that for low back pain also revealed cost-effectiveness in a relatively short time horizon (12 weeks to 3 years) without considering mortality rates.^13–15^ Although the DTx usage half-time was around 15 months in this model, the benefit of the BP-lowering effect persisted for long life above 3 years, resulting in prevention of the trajectory of CVD from hypertension, left ventricular hypertrophy, atherosclerotic events, and finally to HF and CVD deaths. Therefore, a long-term cost-effectiveness analysis would be valuable when considering the practical effects of DTx for chronic diseases such as hypertension.

The monthly cost of DTx for hypertension had the greatest impact on ICERs in the one-way deterministic sensitivity analysis. Lewkowicz et al. pointed out the similar conclusion by evaluating the cost-effectiveness of DTx for low back pain.^15^ At a DTx cost of ¥5,000/month, DTx dominated conventional therapy, or being less costly and more effective option. Even when we set the cost at ¥20,000/month, the ICER was approximately ¥3.5 million/QALY, still under the threshold value of ¥5 million/QALY (**Figure 2**). Recently, sacubitril/valsartan (angiotensin receptor-neprilysin inhibitor), a novel antihypertensive drug, was approved for hypertension management in Japan^34^, with a medication cost of ¥6,057 ($52.67) to ¥12,114 ($105.34) /month. Thus, the DTx cost we set around ¥10,000/month as the default might be a reasonable value according to the current simulation model.

The attrition rate of the DTx program is a crucial factor for cost-effectiveness. A smartphone mHealth app usage rate was attenuating over time, reported approximately 40% of attrition occurred in 6 months to 1 year.^35^ Previous DTx studies for cost-effectiveness analyses also used similar attrition rates.^15, 32^ Thus, it could be reasonable to set the attrition rate of 25% every 6 months in this model, which half time was around 15 months since the initiation of DTx. In another scenario, when we changed the rate to 10% (lower than the predefined 25%), the half time exceeded approximately 40 months, and the ICER was increased to approximately ¥4.3 million/QALY in a lifetime horizon, which was close to the threshold value (**Figure 2**). Lewkowicz et al. previously reported that lowering the DTx app attrition rate was essential for cost-effectiveness in a relatively short-term (3 years) horizon.^15^ Although the model inputs and target disease were different, it might be possible that high attrition rates along with DTx actual usage duration could substantially impact DTx cost-effectiveness. Achieving good cost-effectiveness in DTx, we might face sensitive handling to balance the appropriate DTx app usage duration with DTx costs and expected attrition rate.

The degree of the gained ICER in DTx hypertension management surely depends on the degree of BP reduction by quality of the DTx app algorithm, the impact of BP reduction, and the cost of cardiovascular events in each country. A significant difference in home BP reduction (−4.3 mmHg of morning home SBP) was clearly found between the DTx+TAU and TAU-only groups from the previous phase III clinical trial. It is noteworthy that this TAU-only group followed the ideal guideline-driven management of hypertension, which used home BP monitoring.^7^ Compared with office BP, home BP is more closely associated with cardiovascular event risk.^36, 37^ In addition, in Asian countries, the benefit of BP lowering is greater than that of Westerners, especially for stroke.^38^ The comparison of cost-effectiveness studies using different DTx apps in different contigs is also needed in the future.

The strength of this study is that, to our knowledge, this is the first study to evaluate the cost-effectiveness of DTx for patients with hypertension using the results from an RCT. Furthermore, we demonstrated that DTx+TAU is cost-effective for patients with hypertension. This study has some limitations. First, we simulated middle-aged, digital-friendly grade I or II hypertensive patients without taking antihypertensive medication from the previous RCT results. Thus, caution should be taken when applying our results to a broader population. Second, we modelled the DTx effect as permanent, although the effect estimates were derived from the 6-months duration of the HERB-DH1 trial results. Although we set the BP reduction effects even for the TAU-only strategy, this could lower the mortality and complication occurrence rates in the DTx+TAU strategy. Third, we did not estimate additive effects of having multiple comorbidities in this model, which could impact lifetime ICER. Fourth, we could not strictly assess the DTx cost-effectiveness in addition to antihypertensive medications since the model patients derived from previous trials were all antihypertensive drug-naïve. Finally, we did not consider re-using DTx after finishing the initial DTx usage according to the attrition rate, which could account for additional costs for the DTx strategy.

In conclusion, DTx+TAU demonstrated good cost-effectiveness in hypertensive patients compared to the TAU-only strategy. DTx cost and its attrition rate largely influenced cost-effectiveness, and we need to explore the balance among the DTx application cost, app attrition rate, and effectiveness for target hypertensive patients in clinical practice. DTx is generally reimbursed by Japan’s national universal health insurance system. Thus, it is becoming increasingly essential to evaluate the cost-effectiveness of DTx, as in this study, to consider the appropriate value-based reimbursement pricing.

## Data Availability

All data in the present study are available from the corresponding author upon reasonable request.

## Acknowledgments

We would like to express our gratitude to all participants regarding this study.

