## Supplemental material for "Cost-effectiveness of digital therapeutics for essential hypertension"

Akihiro Nomura, Tomoyuki Tanigawa, Kazuomi Kario, Ataru Igarashi

**Supplemental Table.** Hazard ratios of event occurrence for each complication by BP grades

| Data input | Normal BP | Elevated BP | Grade I HT | Grade II HT | Source |
|---|---|---|---|---|---|
| Acute coronary syndrome | 1 (ref) | 1.14 | 1.46 | 1.96 | Kaneko, 2021.[1] |
| Stroke | 1 (ref) | 1.13 | 1.35 | 2.14 | Kaneko, 2021.[1] |
| Heart failure | 1 (ref) | 1.1 | 1.3 | 2.05 | Kaneko, 2021.[1] |
| Atrial fibrillation | 1 (ref) | 1.07 | 1.21 | 1.52 | Kaneko, 2021.[1] |

| Data input | Elevated BP | Low* | High* |
|---|---|---|---|
| Acute coronary syndrome | 1.14 | 1.01 | 1.30 |
| Stroke | 1.13 | 1.06 | 1.21 |
| Heart failure | 1.1 | 1.05 | 1.15 |
| Atrial fibrillation | 1.07 | 0.99 | 1.17 |

* Range for sensitivity analysis. We used only the elevated BP range values for the sensitivity analysis.

Abbreviations: BP, blood pressure; HT, hypertension

**Supplemental Figure.** Distribution of treatment effects for SBP obtained from the HERB-DH1 pivotal trial.[2] A: DTx + TAU group. B: TAU-only group

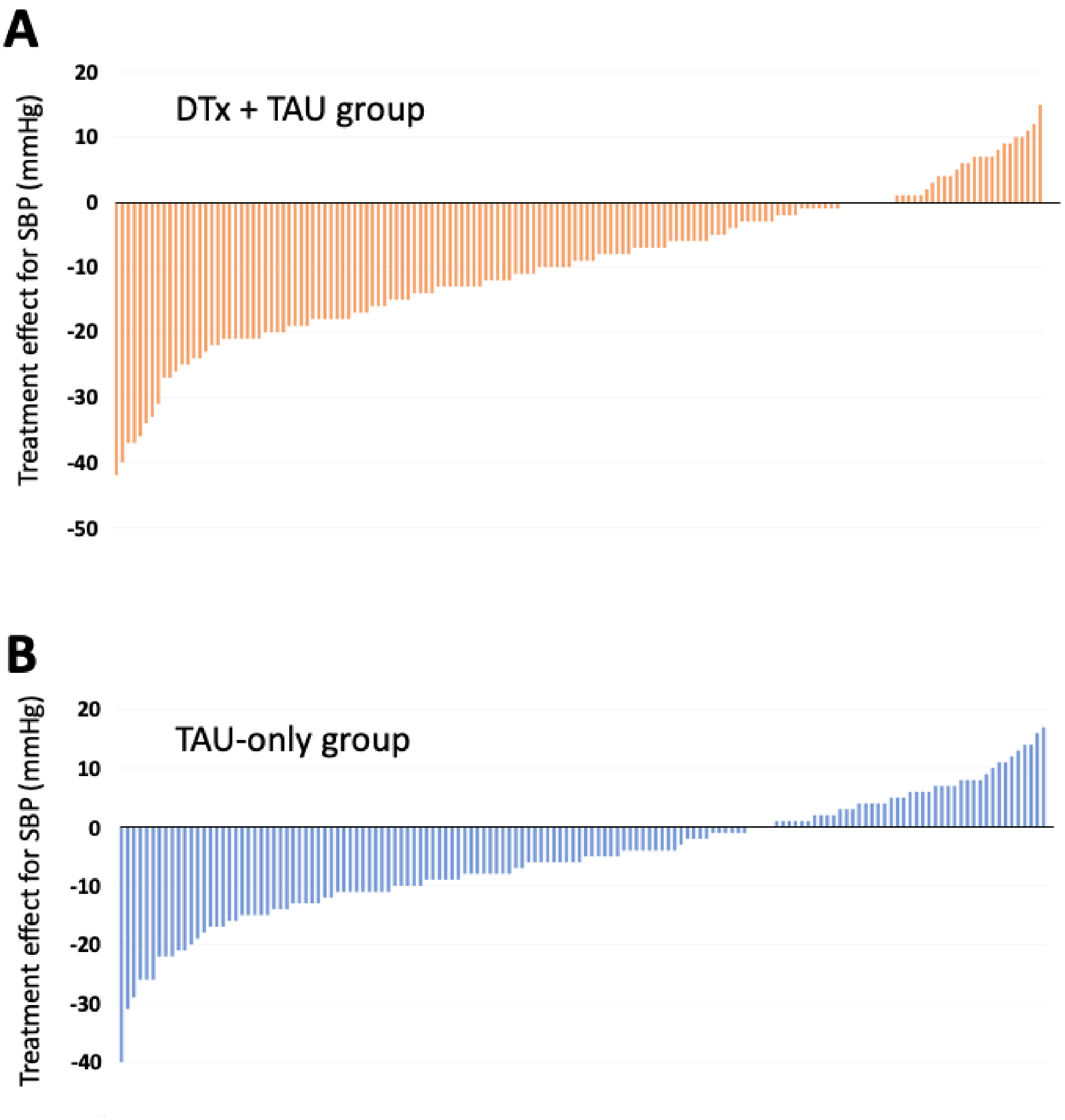
